# Characterization of Trajectories of Physical Activity and Cigarette Smoking from Early Adolescence to Adulthood

**DOI:** 10.1101/2022.11.15.22282238

**Authors:** Iris Yuefan Shao, Shakira F. Suglia, Weihua An, David Mendez, Viola Vaccarino, Alvaro Alonso

## Abstract

**Background:** Cigarette smoking and physical inactivity are two critical risk factors for noncommunicable diseases and all-cause mortality. However, few studies have compared the long-term trajectories of both behaviors, as well as multilevel factors associated with trajectory patterns. Using the National Longitudinal Study of Adolescent to Adult Health (Add Health) Wave I through V survey data, this study characterized distinct subgroups of the population sharing similar patterns of physical activity (PA) and cigarette smoking from adolescence to adulthood, as well as predictors of subgroup membership.

**Methods:** Using the Add Health Wave I through V survey data, we identified the optimal number of latent classes and class-specific trajectories of PA and cigarette smoking from early adolescence to adulthood, fitting latent growth mixture models with standardized PA score and past 30-day cigarette smoking intensity as outcome measures and age as a continuous time variable. Associations of baseline sociodemographic factors, neighborhood characteristics, and sociopsychological factors with trajectory class membership were assessed using multinomial logistic regression.

**Results:** We identified three distinct subgroups of PA trajectories in the study population: moderately active group (N=1067, 5%), persistently inactive group (N=14257, 69%) and worsening activity group (N=5410, 26%). Similarly for cigarette smoking, we identified three distinct trajectory subgroups: persistent non-smoker (N=14939, 72%), gradual quitter (N=2357, 11%), and progressing smoker (N=3393, 16%). Sex, race/ethnicity, neighborhood environment and perceived peer support during adolescence were significant predictors of physical activity and cigarette smoking trajectory subgroup membership from early adolescence to adulthood.

**Conclusion:** There are three distinct subgroups of individuals sharing similar both PA and cigarette smoking behavioral profile from adolescence to adulthood in the Add Health study population. Modifiable risk factors such as neighborhood environment and relationship to peers during adolescence can be key to designing effective behavioral interventions for long-term PA promotion and cigarette smoking cessation.

## Introduction

Cigarette smoking and physical inactivity have long been identified as two critical behavioral risk factors for cardiometabolic diseases and all-cause mortality.^1^ As two modifiable health behaviors, despite their distinct nature, physical activity (PA) and cigarette smoking behavior have been shown to share various similarities.^2–5^ From a theoretical perspective, Social Cognitive Theory^6,7,8^ and the Social Ecological Model^9–11^ have been the two most widely applied theories in explaining patterns associated with both behaviors. Based on these two theories, cognitive factors (e.g. belief), behavioral factors (e.g. self-efficacy) and environmental factors (e.g. social support) are associated with initiation and maintenance of both behaviors. As a result, similar types of behavioral interventions have been proposed. For example, workplace and community support programs have been established for both physical activity promotion and tobacco cessation.^12,13^ Nevertheless, results from similar types of intervention trials targeting PA and cigarette smoking have been mixed.^14,15,16,17^ Therefore, it is important to empirically compare and contrast contributing factors of long-term patterns of these two health behaviors in order to better understand why certain interventions fail at targeting one behavior versus the other.

To better intervene upon these two modifiable health behaviors, exploratory studies have been put forth in recent years to identify long-term patterns of cigarette smoking and physical activity behaviors respectively in heterogenous population.^18–27^ However, most analyses characterizing trajectories of the behaviors have used latent class growth analysis (LCGA), which pre-assumes all latent classes identified are drawn from a single population.^28^ This approach often fails to capture distinct behavior patterns and to take into consideration unobserved distinct subgroups within a population, which is vital to the design and evaluation of person-centered behavioral interventions. Latent Class (Growth) Mixture Models (LCMM) allow for exploration of number and characteristics of unobserved population subgroups that share similar behavior patterns.^28,29^ Using the National Longitudinal Study of Adolescent to Adult Health (Add Health) Wave I through V survey data, this study aims to identify distinct characteristics of subgroup population sharing similar patterns of PA and cigarette smoking from early adolescence to adulthood, as well as predictors of subgroup membership.

## Methods

### Study Design

The Add Health study is a longitudinal cohort study that enrolled a nationally representative sample of adolescents in the United States between grades 7 and 12 at baseline.^30^ It was originally designed to facilitate a multidisciplinary approach to better understand causes of adolescent health behavior and outcomes throughout multiple developmental phases. At baseline (Wave I, 1993-1994), 20,745 participants completed an in-school interview or at-home interview with a mean age of 15 years old. In addition, participants’ parents were invited to complete interviews regarding parental sociodemographic background and household-level socioeconomic information. Four additional waves of data were collected subsequently. Across all five waves, the following information was collected: participants’ socio-demographic information, school performance, peer relationship, biomarker information, health outcomes, health behaviors, romantic relationship, familial and neighborhood-level socio-environmental contextual information, and geospatial information. The present analysis used the in-school questionnaire, parental interview questionnaire and in-person interview questionnaire of the Add Health study from Wave I to Wave V. Eligible study participants have completed either PA or cigarette smoking-related questionnaire items and individuals with missing information across all five waves were excluded the analysis. The use of the data was reviewed and approved by the Institutional Review Board at Emory University and the Add Health study review boards.

### Cigarette Smoking

Survey respondents were asked to self-report cigarette smoking behaviors during in-school and in-home interviews.^31^ Questions regarding lifetime history of cigarette smoking and past 30-day (p30-day) cigarette smoking behavior were asked. In Wave I and II, the following questions were asked to determine respondents’ current smoking status and p30-day cigarette smoking intensity: 1) Have you ever tried cigarette smoking, even just 1 or 2 puffs? 2) Have you ever smoked cigarettes regularly, that is, at least 1 cigarette every day for 30 days? 3) During the past 30 days, on how many days did you smoke cigarettes? 4) During the past 30 days, on the days you smoked, on average, how many cigarettes per day did you smoke? In Wave III through V, the following questions were asked: 1) Have you ever tried cigarette smoking, even just one or two puffs? 2) Have you ever smoked an entire cigarette? 3) Have you smoked at all in the past 30 days? 4) During the past 30 days, on how many days did you smoke cigarettes? 5). During the past 30 days, on the days you smoked, on average, how many cigarettes per day did you smoke? Based on these sets of questions, a dichotomous variable categorizing respondents as current smoker and current non-smoker was generated for all waves. Current smokers were defined as those that have tried cigarettes and smoked cigarettes in the past 30 days. P30-day cigarette smoking intensity was defined as total number of cigarettes smoked in the past 30 days. If respondent was categorized as current non-smoker, p30-day cigarette smoking intensity was zero. Otherwise, p30-day cigarette smoking intensity was calculated as the product of number of days smoked in the past 30 days and number of cigarettes smoked on average on the days respondents smoked. In addition, whether smokers were present in the household during baseline visit was reported as a binary response.

### Physical Activity

Study respondents were asked to self-report how often (times per week) they were engaged in a series of standard physical activities including jogging, walking, karate, jumping rope, gymnastics, dancing, roller-blading, roller-skating, skate-boarding, bicycling, or active sports.^31^ Previous studies^32,33^ have frequently used the definition of moderate-vigorous leisure-time physical activity through approximating number of metabolic equivalents. In this study, instead of using number of metabolic equivalents approximated, we generated a physical activity score corresponding to self-reported physical activity frequency of each questionnaire item to account for change in reported activity categorization in Wave V. If frequency was zero in the past seven days, then the score was assigned as zero. If frequency was either once or twice in the past seven days, then the score was assigned as 1.5. Otherwise a score of 3.5 was assigned. A summary physical activity score was generated by summing up physical activity scores across all questionnaire items at each wave. Additionally, a standardized physical activity score across all five waves was generated by dividing the summary score by number of activities included in each wave’s questionnaire to account for changes in Add Health questionnaire design starting from Wave III.^31^

### Other Variables of Interest

#### Baseline Sociodemographic Characteristics

Sociodemographic variables of interest included biological sex, race/ethnicity, parental education and household income reported at baseline visit.^31^ Survey respondents self-identified as White, African American, Hispanic, Asian/Pacific Islander/Native American/Alaska Indians, or Others. 83% (N = 17238) respondents’ parents participated in the baseline parental interview questionnaire in 1994.^31^ Highest level of parental education obtained by 1994 was reported. Respondents’ parents were further dichotomized as having received a degree no more than high school or having received a degree beyond high school. In addition, total household pre-tax income including welfare benefits, dividends, and others was reported. A four-level ordinal variable was generated based on quartiles of reported household income.

#### Baseline Neighborhood Social Environment

Respondents’ closeness with people in the neighborhood was captured by asking whether they knew most people in the neighborhood. In addition, all respondents to the in-home interview were asked about whether they were happy with the present neighborhood, whether they felt safe in the current neighborhood and whether they had access to a fitness or recreational center in the neighborhood. ^31^

#### Baseline Sociopsychological Factors

Perceived parental, peer and teachers’ support was captured during baseline in-home interviews through questions on whether respondents felt cared for by adults, teachers, and friends. Whether respondents perceived being part of the school or close to others at school was also recorded as a binary response in in-school questionnaires at baseline.^31^

#### Statistical Analysis

Participants who participated in Wave I through Wave V in-school and in-home interviews as well as baseline parental interview questionnaire of the Add Health study with non-missing information on age, PA and cigarette smoking behaviors were included in the analyses. To ensure participants in the five waves of the Add Health study were comparable, key sociodemographic characteristics of study participants by wave were assessed (**Table 1**). To identify subgroups of physical activity and cigarette smoking trajectories from young adolescence to adulthood, latent class mixture models (LCMM) were used. LCMM allows for exploration of population-level outcome heterogeneity by identifying the underlying number of latent classes and accounting for individual-level measure heterogeneity.^34,35^

**Table 1.** Characteristics of the Add Health Study Participants across Five Waves of Study Follow-up, 1994 – 2018.

|  | **Wave I** | **Wave II** | **Wave III** | **Wave IV** | **Wave V** |
| --- | --- | --- | --- | --- | --- |
| **N** |  |  |  |  |  |
| Total | 20745 | 14736 | 15197 | 15701 | 12283 |
| **Parental Education** |  |  |  |  |  |
| College or above | 6941 (33) | 5129 (35) | 5278 (35) | 5372 (34) | 4457 (36) |
| Graduated from high school but not college | 4192 (20) | 2938 (20) | 3088 (20) | 3269 (21) | 2565 (21) |
| Not graduated from high school | 8572 (41) | 6070 (41) | 6153 (41) | 6386 (41) | 4764 (39) |
| **Race/Ethnicity** |  |  |  |  |  |
| Non-Hispanic White | 10455 (50) | 7573 (51) | 7864 (52) | 8294 (53) | 6842 (56) |
| Non-Hispanic Black | 4669 (23) | 3244 (22) | 3316 (22) | 3498 (22) | 2473 (20) |
| Hispanic | 3525 (17) | 2487 (17) | 2447 (16) | 2498 (16) | 1825 (15) |
| Non-Hispanic Asian/Pacific Islander/American Indian/Alaska Native | 1467 (7) | 1004 (7) | 1108 (7) | 947 (6) | 793 (6) |
| Other | 629 (3) | 428 (3) | 435 (3) | 437 (3) | 328 (3) |
| **Female** | 10263 (49) | 7182 (49) | 7167 (47) | 7349 (47) | 5324 (43) |
| **Current Smoker** | 5326 (26) | 4648 (32) | 4786 (32) | 5508 (35) | 2984 (24) |
| **Mean (SD)** |  |  |  |  |  |
| Age (years) | 15.7 (1.7) | 16.2 (1.6) | 22.0 (1.8) | 28.5 (1.8) | 37.5 (1.9) |
| Standardized physical activity score | 0.4 (0.2) | 0.4 (0.2) | 0.2 (0.2) | 0.2 (0.2) | 0.1 (0.1) |
| *\*SD: standard deviation* |  |  |  |  |  |

To determine the optimal number of latent classes and class-specific trajectories of physical activity scores from young adolescence to adulthood for Add Health participants, we fit LCMMs with standardized physical activity score as the outcome measure and age as a continuous time variable. Maximum likelihood measures of a single latent class model were used as the initial values for model estimation. For each model with a hypothesized number of latent classes, model fitting and estimation process were iterated over random vectors of initial values through an automatic grid search algorithm until model achieved the best log-likelihood measure.

Moreover, quadratic trajectories of physical activity scores were explored in addition to linear trajectories. Based on prior literature^33,36,37^, we hypothesized that there were three classes of distinct trajectories. Hence, all model fitting and estimation procedures were iterated over two to four hypothesized number of latent classes in LCMM. Posterior probabilities of participants belonging to a class, given the hypothesized number of classes were obtained. Optimal number of classes for physical activity score trajectories was determined based on the following six factors: model entropy, the Akaike Information Criterion (AIC), the Bayesian Information Criterion (BIC), trajectory shape fitting between predicted and observed data, average posterior probability of individuals belonging to the assigned class (ideally greater or equal to 0.7), and proportion of individuals belonging to each class. With respect to trajectories of p30-day cigarette smoking intensity, a similar approach as described above was used. We hypothesized that three classes of trajectories would be observed.^27^ Log of p30-day cigarette smoking intensity was used as the outcome measure for model fitting purposes. To further identify predictors of latent class membership, multinomial logistic regression was used to assess the association of individual-level predictors (e.g. socio-demographic, psychological factors) and community-level predictors (e.g. household and neighborhood factors) with trajectory class membership for both PA and cigarette smoking. Individuals with missing information regarding baseline predictors were excluded from the analysis. All statistical analyses were performed in R 3.5.2.

## Results

Of 20745 baseline study participants, 14736 (71%) participated in Wave II, 15197 (73%) participated in Wave III, 15701 (76%) participated in Wave IV, and 12283 (59%) participated in Wave V. Across all five waves, the proportion of female participants and participants of different race/ethnicity was comparable. Similarly, participants across all waves reported similar levels of parental education levels (**Table 1**). Of all baseline study participants, 20734 had completed at least one set of physical activity-related questions across five waves and 20689 had completed at least one set of cigarette smoking-related questions across five waves. Amongst respondents included in the PA trajectory analyses, 10474 (51%) were female and 10292 (50%) were non-white participants. With respect to cigarette smoking, 10455 (51%) respondents included in the final analyses were female and 10308 (50%) were non-white participants (**Table 2, Table 3**).

**Table 2.** Baseline Class Member Profile of Physical Activity Trajectory.

|  | **Physical Activity Trajectory Class Profile** |  |  |
| --- | --- | --- | --- |
|  | **Class 1**<br>**(Moderately**<br>**active)** | **Class 2**<br>**(Persistently**<br>**inactive)** | **Class 3**<br>**(Worsening**<br>**activity)** |
| N (%) | 1067 (5) | 14257 (69) | 5410 (26) |
| **Socio-demographic** |  |  |  |
| Female | 303 (28) | 8324 (58) | 1847 (34) |
| Non-Hispanic White | 503 (47) | 7059 (50) | 2891 (53) |
| Non-Hispanic Black | 247 (23) | 3348 (23) | 1072 (20) |
| Hispanic | 181 (17) | 2447 (17) | 893 (17) |
| Other | 136 (12) | 1403 (10) | 554 (10) |
| Parental Education (Less than High school) | 388 (36) | 6167 (43) | 2016 (37) |
| Household Income (lowest quartile) | 238 (22) | 4104 (29) | 1324 (24) |
| **Neighborhood** |  |  |  |
| Knowing most people in the neighborhood | 814 (76) | 9577 (67) | 4089 (76) |
| Happy with present neighborhood | 986 (92) | 12754 (89) | 4968 (92) |
| Access to recreational center in neighborhood | 302 (28) | 2360 (17) | 1548 (29) |
| Feel safe in the neighborhood | 968 (91) | 12382 (87) | 4831 (89) |
| **Socio-psychological\*** |  |  |  |
| Perceived adults' care | 1022 (96) | 13657 (96) | 5218 (96) |
| Perceived teachers' care | 924 (87) | 12111 (85) | 4695 (87) |
| Perceived friends' care | 1040 (97) | 13733 (96) | 5236 (97) |
| Perceived closeness with peers at school | 467 (44) | 5168 (26) | 2404 (44) |
| Perceived as part of school | 457 (43) | 5101 (36) | 2357 (44) |
| *Socio-psychological characteristics were coded as dichotomous variables. |  |  |  |

**Table 3.** Baseline Class Member Profile of Past 30-day Cigarette Smoking Intensity Trajectory.

|  | **Past 30-day Cigarette Smoking Trajectory Class Profile** |  |  |
| --- | --- | --- | --- |
|  | **Persistent non-smoker** | **Gradual quitter** | **Progressing smoker** |
| **N (%)** | 14939 (72) | 2357 (11) | 3393 (16) |
| **Socio-demographic** |  |  |  |
| Female | 7800 (52) | 1181 (50) | 1474 (43) |
| Non-Hispanic White | 6691 (45) | 1734 (74) | 2012 (59) |
| Non-Hispanic Black | 3781 (25) | 128 (5) | 742 (22) |
| Hispanic | 2900 (19) | 270 (11) | 342 (10) |
| Other | 1567(11) | 225 (9) | 297 (9) |
| Parental Education (Less than High school) | 5899 (39) | 1095 (46) | 1556 (46) |
| Household Income (lowest quartile) | 3951 (26) | 641 (27) | 1058 (31) |
| Presence of smoker in household | 4822 (32) | 1435 (61) | 1732 (51) |
| **Neighborhood** |  |  |  |
| Knowing most people in the neighborhood | 10187 (68) | 1660 (70) | 2611 (77) |
| Happy with present neighborhood | 13537 (91) | 2093 (89) | 3046 (90) |
| **Socio-psychological\*** |  |  |  |
| Perceived adults' care | 14353 (96) | 2253 (96) | 3254 (96) |
| Perceived teachers' care | 13162 (88) | 1765 (75) | 2774 (82) |
| Perceived friends' care | 14401 (96) | 2302 (98) | 3270 (96) |
| Perceived closeness with peers at school | 6026 (40) | 769 (33) | 1232 (36) |
| Perceived as part of school | 6049 (40) | 671 (28) | 1184 (35) |
| *Socio-psychological characteristics were coded as dichotomous variables. |  |  |  |

### Trajectory Classes and Class Member Profile for Standardized Physical Activity Score

We identified three distinct subgroups of PA trajectories in the study population: moderately active group (Class 1, N= 1067, 5%), persistently inactive group (Class 2, N= 14257, 69%) and worsening activity group (Class 3, N= 5410, 26%) since three classes resulted in the model with the highest entropy, smallest AIC/BIC as well as mean posterior probability of individual truly belonging to each class greater than 0.70 (**Table 2, Supplemental Table 1, and Figure 1**). The moderately active group maintained a moderate PA level till 30 years old, when PA level dropped. The persistently inactive group had the lowest PA level across all groups over time and had the smallest magnitude of change in PA level over time. The worsening activity group had the highest PA level prior to 15 years old but the PA level dropped drastically starting at 15 years old and leveled off starting at 25 years old. The magnitude of change in physical activity level was the biggest in this group. Overall, prior to 18 years of age, the worsening activity group had the highest mean PA level whereas the moderately active group became the most active group amongst all groups starting at 18 years old (**Figure 1**).

**Figure 1.**
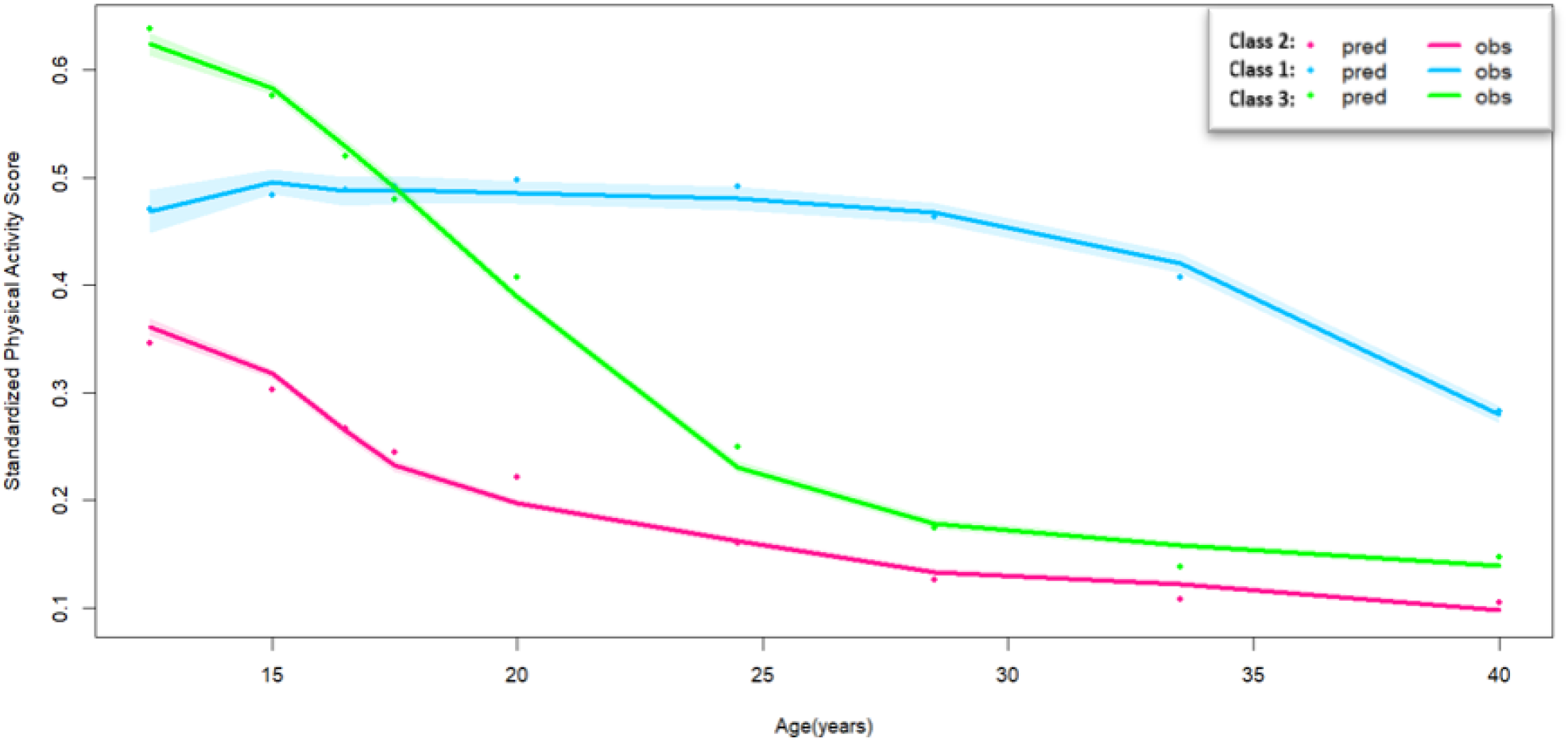
Subject-specific Trajectories of Standardized Physical Activity Score from Early Adolescence to Adulthood.

With respect to baseline socio-demographic characteristics, the persistently inactive group had the highest proportion of females (N = 8324, 58%), parents that did not receive a high school degree or above (N = 6167, 43%), and households with an income in the lowest quartile (N = 4104, 29%) whereas the moderately active group had the lowest proportion of females (N= 303, 28%), the lowest number of parents that did not receive a high school degree or above (N = 388, 36%) and the lowest number of households with an income in the lowest quartile (N = 238, 22%). With respect to baseline neighborhood characteristics, the persistently inactive group had the lowest proportion of respondents that reported being happy with their present neighborhood (N=12754, 89%), had access to a recreational center in the neighborhood (N = 2360, 17%), felt safe in the neighborhood (N = 12382, 87%), or knew almost everyone in the community (N=9577, 67%). Perceived closeness with peers at school and perceiving as a part of the school appeared to be two differentiating factors between the persistently inactive group and the other two groups. Proportions of individuals that perceived as close with peers at school and perceived as part of the school were lowest in the persistently inactive group. (**Table 2**)

### Trajectory Classes and Class Member Profile for Past 30-day Cigarette Smoking Intensity

With respective to past 30-day cigarette smoking intensity, we observed three distinct groups in the study population: persistent non-smoker (Class 1, N=14939, 72%), gradual quitter (Class 2, N=2357, 11%), and progressing smoker (Class 3, N=3393, 16%) since three classes resulted in the model with the highest entropy, smallest AIC/BIC as well as mean posterior probability of individual actually belonging to each class greater than 0.70 (**Table 3, Supplemental Table 1**). The gradual quitter group increased p30-day cigarette smoking intensity prior to 18 years of age and started reducing p30-day cigarette smoking intensity throughout adulthood. The persistent non-smoker group remained as non-smoker throughout the entire study follow-up period. The progressing smoker group increased p30-day cigarette smoking intensity from adolescence to adulthood consistently and had the highest magnitude of increase from adolescence to young adulthood. Rate of increase in p30-day cigarette smoking intensity amongst this group decreased starting at 22 years of age and plateaued around 26 years of age till the end of the study follow-up. The gradual quitter group had the highest mean log (p30-day cigarette smoking intensity) prior to 23 years of age. After 23 years of age, p30-day cigarette smoking intensity amongst the progressing smoker group became the highest (**Figure 2**).

**Figure 2.**
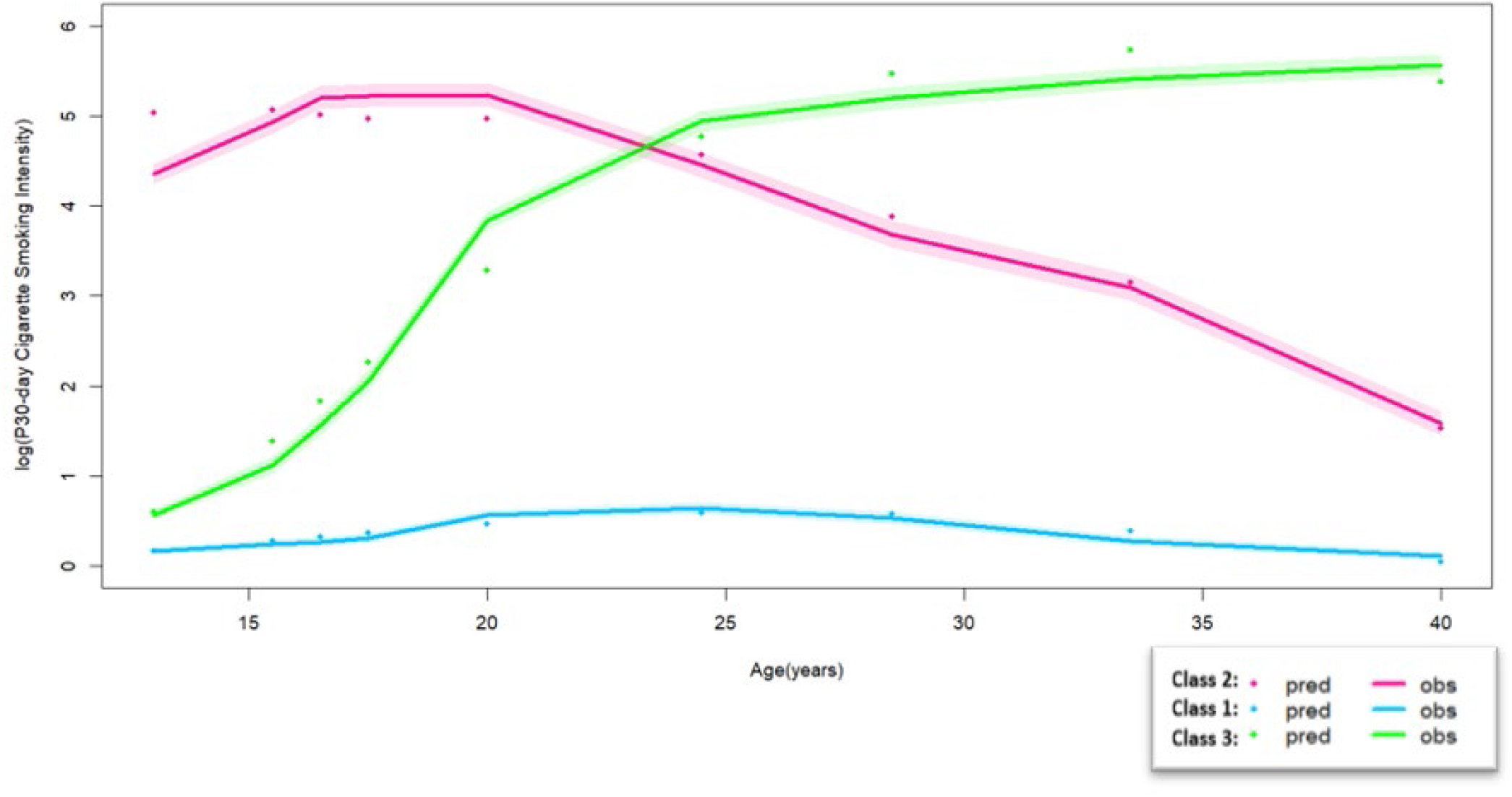
Subject-specific Trajectories of Log (Past 30-day Cigarette Smoking Intensity) from Early Adolescence to Adulthood.

Among the three groups, the progressing smoker group had the lowest proportion of females (N = 1474, 43%), meanwhile the persistent non-smoker group had the highest (N = 7800, 52%). The gradual quitter group had the lowest proportion of racially disadvantaged population (N = 623, 26%). The gradual quitter group had the most individuals indicating the presence of smokers in the household (N = 1435, 61%) whereas the persistent non-smoker had the fewest. (N = 4822, 32%). Interestingly, the persistent non-smoker group had the fewest respondents indicating knowing most people in the neighborhood (N = 10187, 68%), whereas the progressing smoker group had the most (N = 2611, 77%). With respect to baseline socio-psychological factors, perceived closeness with peers at school, perceived care from teachers, and perceived as part of the school appeared to be three differentiating factors. The persistent non-smoker group had the highest proportions of individuals that responded positively to all questions above whereas the gradual quitter group had the lowest proportions (**Table 3**).

### Predictors of Physical Activity and Past 30-day Cigarette Smoking Intensity Class Membership

Based on results from multinomial logistic regression analyses, being male, higher baseline parental education and parental income, having access to fitness center in the community, perceived happiness and closeness in the neighborhood, perceived closeness and sense of belonging at school were significant predictors of being in the moderately active group as compared to the persistently inactive group. On the contrary, being female, being non-Hispanic white, and lower baseline parental income appear to be the only significant predictors of being in the worsening activity group as compared to being in the moderately active group (**Table 4**). Overall, females were more likely to be in the persistently inactive and worsening activity groups (RRR = 3.59, 95% CI: 2.97-4.33; RRR = 1.47, 95% CI: 1.20-1.78). Baseline neighborhood characteristics only differentiated individuals in the persistently inactive group versus the moderately active group. In addition, not feeling as part of school is a significant predictor of an individual being persistently inactive (RRR= 0.72, 95% CI: 0.53-0.96).

**Table 4.**
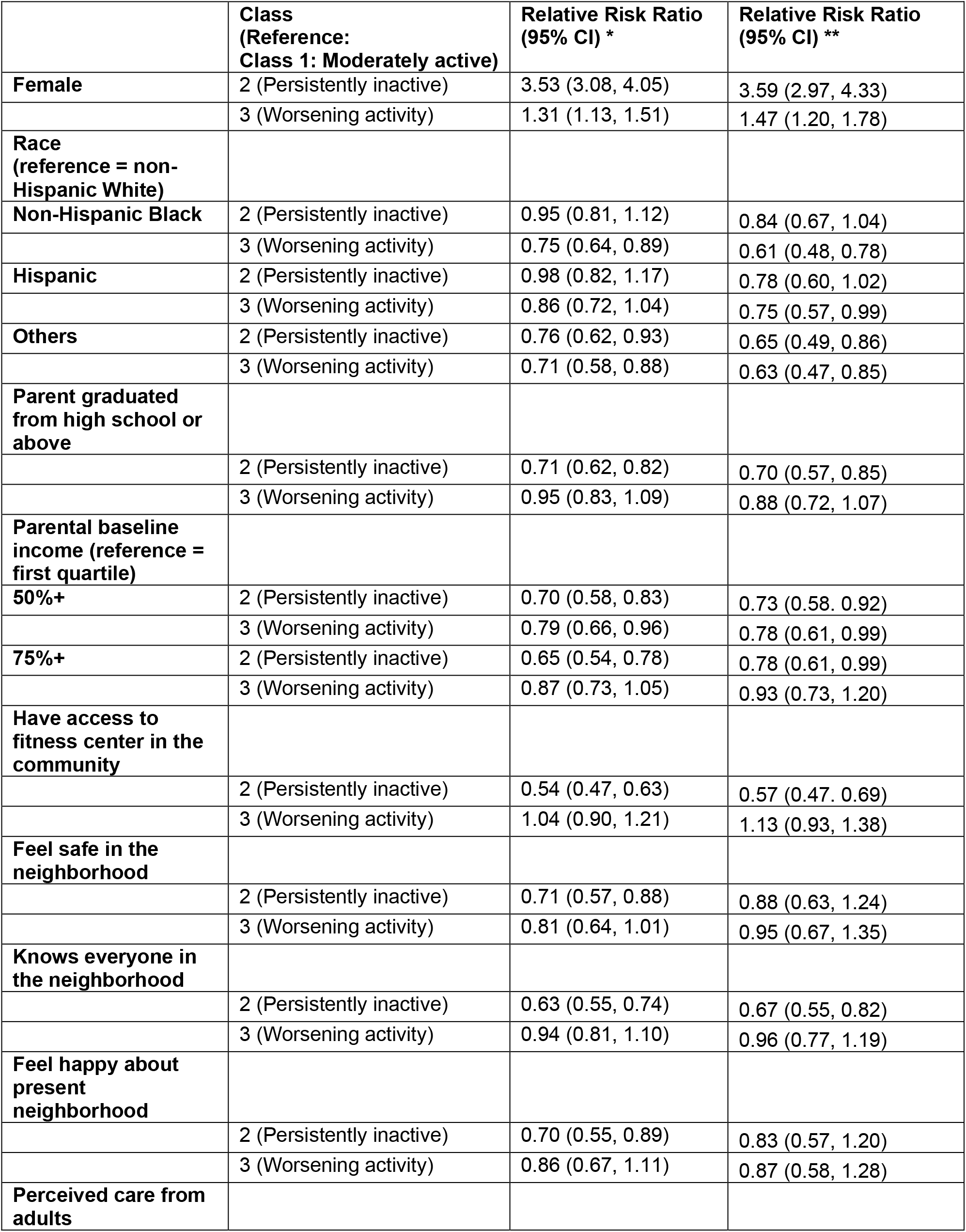
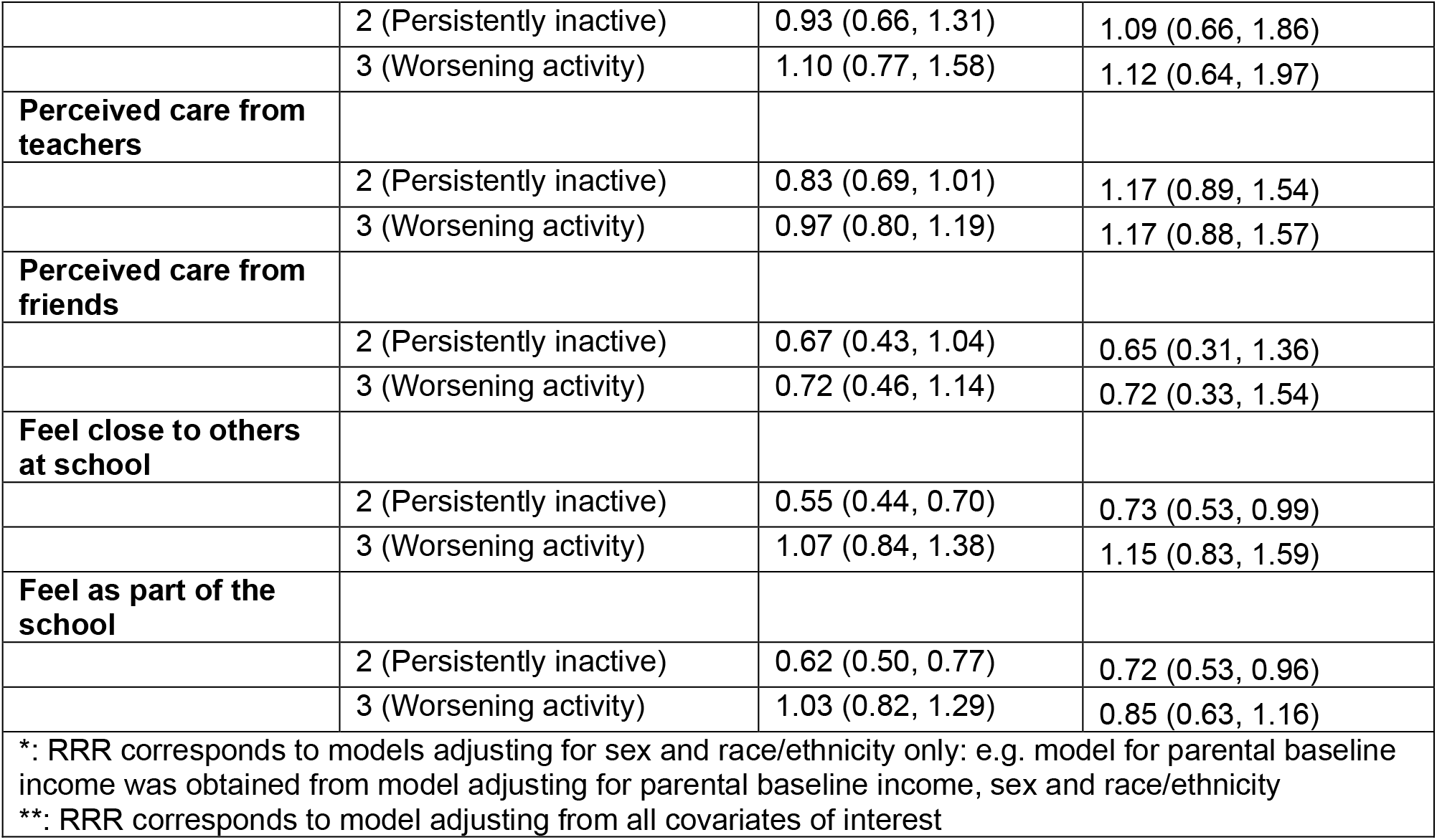
Predictors of Physical Activity Trajectory Class Membership.

|  | **Class**<br>**(Reference:**<br>**Class 1: Moderately active)** | **Relative Risk Ratio**<br>**(95% CI) \*** | **Relative Risk Ratio**<br>**(95% CI) \*\*** |
| --- | --- | --- | --- |
| **Female** | 2 (Persistently inactive) | 3.53 (3.08, 4.05) | 3.59 (2.97, 4.33) |
|  | 3 (Worsening activity) | 1.31 (1.13, 1.51) | 1.47 (1.20, 1.78) |
| **Race**<br>**(reference = non-**<br>**Hispanic White)** |  |  |  |
| **Non-Hispanic Black** | 2 (Persistently inactive) | 0.95 (0.81, 1.12) | 0.84 (0.67, 1.04) |
|  | 3 (Worsening activity) | 0.75 (0.64, 0.89) | 0.61 (0.48, 0.78) |
| **Hispanic** | 2 (Persistently inactive) | 0.98 (0.82, 1.17) | 0.78 (0.60, 1.02) |
|  | 3 (Worsening activity) | 0.86 (0.72, 1.04) | 0.75 (0.57, 0.99) |
| **Others** | 2 (Persistently inactive) | 0.76 (0.62, 0.93) | 0.65 (0.49, 0.86) |
|  | 3 (Worsening activity) | 0.71 (0.58, 0.88) | 0.63 (0.47, 0.85) |
| **Parent graduated**<br>**from high school or**<br>**above** |  |  |  |
|  | 2 (Persistently inactive) | 0.71 (0.62, 0.82) | 0.70 (0.57, 0.85) |
|  | 3 (Worsening activity) | 0.95 (0.83, 1.09) | 0.88 (0.72, 1.07) |
| **Parental baseline**<br>**income (reference =**<br>**first quartile)** |  |  |  |
| **50%+** | 2 (Persistently inactive) | 0.70 (0.58, 0.83) | 0.73 (0.58, 0.92) |
|  | 3 (Worsening activity) | 0.79 (0.66, 0.96) | 0.78 (0.61, 0.99) |
| **75%+** | 2 (Persistently inactive) | 0.65 (0.54, 0.78) | 0.78 (0.61, 0.99) |
|  | 3 (Worsening activity) | 0.87 (0.73, 1.05) | 0.93 (0.73, 1.20) |
| **Have access to**<br>**fitness center in the**<br>**community** |  |  |  |
|  | 2 (Persistently inactive) | 0.54 (0.47, 0.63) | 0.57 (0.47, 0.69) |
|  | 3 (Worsening activity) | 1.04 (0.90, 1.21) | 1.13 (0.93, 1.38) |
| **Feel safe in the**<br>**neighborhood** |  |  |  |
|  | 2 (Persistently inactive) | 0.71 (0.57, 0.88) | 0.88 (0.63, 1.24) |
|  | 3 (Worsening activity) | 0.81 (0.64, 1.01) | 0.95 (0.67, 1.35) |
| **Knows everyone in**<br>**the neighborhood** |  |  |  |
|  | 2 (Persistently inactive) | 0.63 (0.55, 0.74) | 0.67 (0.55, 0.82) |
|  | 3 (Worsening activity) | 0.94 (0.81, 1.10) | 0.96 (0.77, 1.19) |
| **Feel happy about**<br>**present**<br>**neighborhood** |  |  |  |
|  | 2 (Persistently inactive) | 0.70 (0.55, 0.89) | 0.83 (0.57, 1.20) |
|  | 3 (Worsening activity) | 0.86 (0.67, 1.11) | 0.87 (0.58, 1.28) |
| **Perceived care from**<br>**adults** |  |  |  |
|  | 2 (Persistently inactive) | 0.93 (0.66, 1.31) | 1.09 (0.66, 1.86) |
|  | 3 (Worsening activity) | 1.10 (0.77, 1.58) | 1.12 (0.64, 1.97) |
| **Perceived care from teachers** |  |  |  |
|  | 2 (Persistently inactive) | 0.83 (0.69, 1.01) | 1.17 (0.89, 1.54) |
|  | 3 (Worsening activity) | 0.97 (0.80, 1.19) | 1.17 (0.88, 1.57) |
| **Perceived care from friends** |  |  |  |
|  | 2 (Persistently inactive) | 0.67 (0.43, 1.04) | 0.65 (0.31, 1.36) |
|  | 3 (Worsening activity) | 0.72 (0.46, 1.14) | 0.72 (0.33, 1.54) |
| **Feel close to others at school** |  |  |  |
|  | 2 (Persistently inactive) | 0.55 (0.44, 0.70) | 0.73 (0.53, 0.99) |
|  | 3 (Worsening activity) | 1.07 (0.84, 1.38) | 1.15 (0.83, 1.59) |
| **Feel as part of the school** |  |  |  |
|  | 2 (Persistently inactive) | 0.62 (0.50, 0.77) | 0.72 (0.53, 0.96) |
|  | 3 (Worsening activity) | 1.03 (0.82, 1.29) | 0.85 (0.63, 1.16) |
| *: RRR corresponds to models adjusting for sex and race/ethnicity only: e.g. model for parental baseline income was obtained from model adjusting for parental baseline income, sex and race/ethnicity |  |  |  |
| **: RRR corresponds to model adjusting from all covariates of interest |  |  |  |

With respect to cigarette smoking, females were less likely to be a progressing smoker as compared to being a gradual quitter (RRR = 0.74, 95% CI: 0.64, 0.86) (**Table 5**). Race was a significant predictor of being a gradual quitter as compared to the other two smoking trajectory groups. Non-Hispanic Black individuals were more likely to be persistent non-smokers (RRR = 7.32, 95% CI: 5.54, 9.66) and progressing smokers (RRR = 4.86, 95% CI: 3.61, 6.53) as compared to gradual quitter. Individuals that reported feeling happy about present neighborhood, perceived care from adults and teachers at baseline, feeling close to others and belonged to the school were significantly more likely to be persistent non-smokers. Meanwhile, presence of a smoker in the household was associated with higher likelihood of being gradual quitter or progressing smoker as compared to persistent non-smoker. Interestingly, individuals that reported feeling close to others and feeling as part of the school were more likely to be a progressing smoker as compared to be a gradual quitter (**Table 5**).

**Table 5.** Predictors of P30-day Cigarette Smoking Intensity Trajectory Class Membership.

|  | **Class**<br>**(Reference:**<br>**Class 2: Gradual quitter)** | **Relative Risk**<br>**Ratio (95% CI) \*** | **Relative Risk Ratio**<br>**(95% CI) \*\*** |
| --- | --- | --- | --- |
| **Female** | 1 (Persistent Non-smoker) | 1.08 (0.99, 1.18) | 1.07 (0.94, 1.21) |
|  | 3 (Progressing Smoker) | 0.75 (0.68, 0.84) | 0.74 (0.64, 0.86) |
| **Race**<br>**(reference = White)** |  |  |  |
| **Non-Hispanic Black** | 1 (Persistent Non-smoker) | 7.64 (6.36, 9.19) | 7.32 (5.54, 9.66) |
|  | 3 (Progressing Smoker) | 5.02 (4.12, 6.12) | 4.86 (3.61, 6.53) |
| **Hispanic** | 1 (Persistent Non-smoker) | 2.79 (2.43, 3.19) | 2.21 (1.80, 2.71) |
|  | 3 (Progressing Smoker) | 1.09 (0.91, 1.29) | 0.97 (0.75, 1.25) |
| **Others** | 1 (Persistent Non-smoker) | 1.81 (1.56, 2.10) | 1.70 (1.33, 2.17) |
|  | 3 (Progressing Smoker) | 1.13 (0.94, 1.36) | 1.43 (1.00, 1.32) |
| **Parent graduated from**<br>**high school or above** |  |  |  |
|  | 1 (Persistent Non-smoker) | 1.58 (1.44, 1.73) | 1.15 (1.00, 1.32) |
|  | 3 (Progressing Smoker) | 1.04 (0.94, 1.17) | 0.93 (0.79, 1.09) |
| **Parental baseline**<br>**income (reference =**<br>**first quartile)** |  |  |  |
| **50%+** | 1 (Persistent Non-smoker) | 1.29 (1.14, 1.45) | 0.98 (0.83, 1.16) |
|  | 3 (Progressing Smoker) | 0.99 (0.86, 1.14) | 0.89 (0.74, 1.08) |
| **75%+** | 1 (Persistent Non-smoker) | 1.52 (1.35, 1.72) | 0.99 (0.83, 1.18) |
|  | 3 (Progressing Smoker) | 0.93 (0.80, 1.07) | 0.75 (0.61, 0.91) |
| **Presence of smoker in**<br>**the household** |  |  |  |
|  | 1 (Persistent Non-smoker) | 0.25 (0.22, 0.28) | 0.30 (0.27, 0.35) |
|  | 3 (Progressing Smoker) | 0.58 (0.51, 0.65) | 0.61 (0.52, 0.72) |
| **Knows everyone in the**<br>**neighborhood** |  |  |  |
|  | 1 (Persistent Non-smoker) | 0.90 (0.82, 0.99) | 0.83 (0.71, 0.96) |
|  | 3 (Progressing Smoker) | 1.33 (1.18, 1.50) | 1.22 (1.02, 1.46) |
| **Feel happy about**<br>**present neighborhood** |  |  |  |
|  | 1 (Persistent Non-smoker) | 1.56 (1.35, 1.80) | 1.40 (1.12, 1.76) |
|  | 3 (Progressing Smoker) | 1.25 (1.05, 1.49) | 1.28 (0.98, 1.67) |
| **Perceived care from**<br>**adults** |  |  |  |
|  | 1 (Persistent Non-smoker) | 1.54 (1.22, 1.94) | 1.11 (0.76, 1.62) |
|  | 3 (Progressing Smoker) | 1.34 (1.02, 1.77) | 1.14 (0.73, 1.77) |
| **Perceived care from**<br>**teachers** |  |  |  |
|  | 1 (Persistent Non-smoker) | 2.84 (2.54, 3.18) | 2.29 (1.92, 2.73) |
|  | 3 (Progressing Smoker) | 1.61 (1.41, 1.85) | 1.60 (1.30, 1.96) |
| **Perceived care from friends** |  |  |  |
|  | 1 (Persistent Non-smoker) | 0.99 (0.72, 1.37) | 0.75 (0.45, 1.26) |
|  | 3 (Progressing Smoker) | 0.82 (0.57, 1.17) | 0.69 (0.39, 1.23) |
| **Feel close to others at school** |  |  |  |
|  | 1 (Persistent Non-smoker) | 1.78 (1.54, 2.06) | 1.03 (0.84, 1.27) |
|  | 3 (Progressing Smoker) | 1.35 (1.14, 1.61) | 1.01 (0.79, 1.28) |
| **Feel as part of the school** |  |  |  |
|  | 1 (Persistent Non-smoker) | 2.35 (2.05, 2.70) | 1.82 (1.50, 2.22) |
|  | 3 (Progressing Smoker) | 1.57 (1.34, 1.86) | 1.32 (1.05, 1.65) |
| *: RRR corresponds to models adjusting for sex and race/ethnicity only: e.g. model for parental baseline income was obtained from model adjusting for parental baseline income, sex and race/ethnicity |  |  |  |
| **: RRR corresponds to model adjusting from all covariates of interest |  |  |  |

## Discussion

This study explored subgroups of individuals sharing similar trajectories of physical activity and past 30-day cigarette smoking behavior from adolescence to adulthood, as well as predictors of specific subgroup membership. Our study revealed three distinct groups of individuals following similar patterns of physical activity (moderately active, persistently inactive, worsening activity) and three distinct groups of individuals following similar patterns of cigarette smoking behavior in the past 30 days (persistent non-smoker, gradual quitter, progressing smoker). In general, physical activity level decreased from adolescence to adulthood. However, cigarette smoking behavioral patterns differed significantly across three groups from adolescence to adulthood. Interestingly, for both physical activity and cigarette smoking behavior, there was one group of individuals that had a consistent behavioral pattern throughout the entire study follow-up and the population size of these groups were the highest. Additionally, transition from adolescence to young adulthood and late adulthood both appeared to be critical to altering individuals’ physical activity patterns whereas transition from young adulthood to late adulthood might be a critical time window for change in cigarette smoking behavior.

These findings are consistent with several earlier studies that also found 3 to 4 subgroups of trajectories for both PA and cigarette smoking.^27,33,38,39,40^ Similar to our study, one existing study on life-course trajectories of PA based on a Finnish sample have shown that overall PA level declines overtime and persistently low-activity group makes up the large proportion of the study population. Major changes in PA level also started during the transition period between adolescence to young adulthood around 21 years old.^40^ Contrastingly, one large study in a US sample described 10 subgroups of trajectories, among which there were additional groups of individuals that remained persistently at high levels of PA, as well as increasingly levels of PA from adolescence to late adulthood.^41^ For cigarette smoking, very few studies explored the long-term trajectories of cigarette smoking behavior spanning across adolescence to adulthood.^27^ One recent study based on a Northern Finnish study population with a 46-year follow-up characterized six different groups of individuals sharing similar patterns of behavior overtime. However, the overall patterns of these six trajectories are similar to findings from our study, indicating the presence of progressing smoker, never smoker and gradual quitter (labeled as quitters in the study).^42^

Our study showed that sex and race/ethnicity are significant socio-demographic predictors of long-term trajectories of PA level and cigarette smoking. Meanwhile, baseline parental education and parental income were significantly associated with trajectory class membership of PA. Such associations were not observed for cigarette smoking behavior. The observed sex difference in PA patterns overtime has long been in discussion. Theories and studies have suggested that females are more likely to be more inactive as compared to their male counterparts, potentially due to long-established gender norms, in addition to physiological differences.^43,44^ With our additional analyses on the association between household/neighborhood level factors and PA trajectory, adjusting for socio-demographic predictors mentioned above, it appeared that socio-demographic factors are key predictors of PA trajectory and such observation could be partially explained through downstream household and neighborhood level factors that impact one’s access to physical activity, for example, limited access to recreational centers and safety issue in a low socio-economic status neighborhood. In terms of baseline socio-psychological predictors of PA trajectory, our study found that perceived closeness with peers as well as feeling as part of the school during adolescence was associated with a decreased likelihood of being persistently inactive throughout the life course, even after adjusting for critical socio-demographic factors. The finding suggested that the adolescence period is critical to shaping lifetime physical activity. Significant numbers of empirical research have shown that peer influence plays a role in moderating physical activity behaviors.^45–47^ In addition to existing findings, our study has shown the possibility of the lasting impact of peer influence and perceived normative behavior at younger age over lifelong trajectories of physical activity.

For cigarette smoking, our study showed that females are more likely to be gradual quitter as compared to progressing smoker, which is contrasting to existing studies indicating that it might be more difficult for females to quit smoking once started.^48,49^ Consistent with prior research, results from our study showed that racially disadvantaged individuals are more likely to be progressing smokers rather than gradual quitters, which suggests that racial minorities, once initiated cigarette smoking, are less likely to quit in the long term.^50^ Interestingly, in our study, non-Hispanic White individuals are more likely to be progressing smokers as compared to racially disadvantaged individuals. Previous studies have also echoed such findings. Various studies^51–53^ have shown that African American, Hispanic and American Indian individuals, despite their age of cigarette smoking initiation, are lighter and intermittent smokers as compared to Whites. Different from PA trajectories, we found that household and neighborhood level predictors are important to differentiate persistent non-smokers as compared to progressing smokers/non-smokers even after adjusting for individual socio-demographic factors, suggesting the importance of contextual exposure to cigarette smoking behavior. However, the presence of a smoker in the household is associated with higher risk of being a progressing smoker and gradual quitter as compared to never smoker. This finding is similar to a previous study showing that household smoking is not linearly and positively correlated with cigarette smoking or quitting in the long-term.^54^ With regards to socio-psychological predictors, in contrast with associations with PA trajectory membership, we did not find an association between baseline socio-psychological factors and long-term trajectories of cigarette smoking intensity.

Results from this study have several important indications for the design of behavioral interventions targeting physical activity and cigarette smoking. First, interventions are still much needed for both behaviors given the large proportions of individuals that are persistently inactive and gradual quitters in our study population. Second, when designing interventions targeting PA, individual sociodemographic and sociopsychological factors might be important to take into consideration, especially when considering the sex difference in motivation to engage in PA. One study has shown that for females, individual-based intervention such as positive messaging and campaign has been more effective in the long run than changes in built environment.^43^ For cigarette smoking, however, individual sociodemographic and contextual factors during early adolescence such as household and neighborhood level factors are key. Third, with regards to potential target population characteristics, our analyses indicate that individuals engaging in better PA behavior over time are more likely to be male, coming from a better socio-economic background, and perceived closeness to others in the neighborhood/peers at a younger age whereas for cigarette smoking, males and those perceiving closeness to others in the neighborhood/peers during early adolescence are less likely to gradually quit cigarette smoking. Collectively, these findings showed that different types of behavioral interventions might be needed when targeting PA versus cigarette smoking. In addition, despite a dire need to address disparity in these two behaviors due to inequality and inequity, a careful examination of intervention design prior to implementation is needed due to greater inertia to behavioral change amongst disadvantaged population.

Our study has several strengths. First, to our knowledge, it is one of the first studies that characterized trajectories of physical activity and cigarette smoking behavior from adolescence to adulthood using a comprehensive large nationally presentative longitudinal study. Second, utilizing latent class growth mixture models, our study was able to identify specific trajectories and population subgroups, taking into consideration both group and individual level heterogeneity. Third, a comprehensive exploration of predictors associated with trajectories allowed for investigation of important factors associated with subgroup membership, which is crucial to identify behavioral intervention target population characteristics as well as intervention design strategies. In the meantime, several limitations need to be acknowledged. First, the number of Add Health study participants decreased by Wave V. Approximately 50% of the study participants were part of Wave V of the study. Even though the sociodemographic characteristics of study participants were comparable across all five waves (**Table 1**), individuals that were not part of the study during later waves of study might lead to missingness in outcome data that are not at random, which might result in biased study findings. Second, both physical activity and past 30-day cigarette smoking data were obtained through self-reported survey. However, the design of the questionnaire items regarding those two behaviors was not consistent across all waves, which might lead to measurement error of outcomes. Third, Wave I through III did not have questions on average cigarette smoking intensity on a typical smoking day in addition to the past 30-day daily cigarette smoking intensity. Therefore, this study used past 30-day cigarette smoking intensity as an outcome, which might not be representative of all cigarette smokers’ typical smoking behavior for longer time periods. Lastly, latent class (growth) mixture model is a post-hoc analytical approach that is constrained by parameters imposed on model specification, such as hypothesized number of groups, as well as whether the trajectory would follow a linear, quadratic or cubic pattern. Even though our study explored different model specification, similar research question needs to be explored in other studies to further confirm the research finding.

## Conclusion

To conclude, our study indicates that age, socio-demographic and psychological factors during early adolescence are important predictors of long-term behavioral trajectories for both PA and cigarette smoking behavior. For PA, modifiable factors during early adolescence such as access to fitness center, perceived closeness to others in the neighborhood and school are associated with an increased likelihood for one to have better long-term physical activity behavior. Contrastingly, for cigarette smoking, perceived closeness to others in neighborhood and friends at school during early adolescence were associated with an increased likelihood of being a progressing smoker than being a gradual quitter. Findings suggest that neighborhood and school environment during early adolescence has a differential impact on promoting long-term health benefitting behaviors such as physical activity and cigarette smoking cessation. Future behavioral interventions targeting modifiable risk factors need to take into consideration timing, target population characteristics and the type of health behavior to be effective.

## Data Availability

All data produced in the present study are available upon reasonable request to the authors

## Funding

This research uses data from Add Health, funded by grant P01 HD31921 (Harris) from the Eunice Kennedy Shriver National Institute of Child Health and Human Development (NICHD), with cooperative funding from 23 other federal agencies and foundations. Add Health is currently directed by Robert A. Hummer and funded by the National Institute on Aging cooperative agreements U01 AG071448 (Hummer) and U01AG071450 (Aiello and Hummer) at the University of North Carolina at Chapel Hill. Add Health was designed by J. Richard Udry, Peter S. Bearman, and Kathleen Mullan Harris at the University of North Carolina at Chapel Hill. Dr. Shao was supported by pilot grant from the Center for the Assessment of Tobacco Regulations at the University of Michigan, Ann Arbor. Dr. Alonso was supported by grant K24HL148521.

## List of Abbreviations

Add Health: National Longitudinal Study of Adolescent to Adult Health
PA: Physical Activity
LCGA: Latent Class Growth Analysis
LCMM: Latent Class (Growth) Mixture Models
RRR: Relative Risk Ratio
CI: Confidence Interval

## Notes

Conflict of Interest Statement: The authors have no conflict of interest to disclose.

### Competing Interest Statement

The authors have declared no competing interest.

### Author Declarations

Institutional Review Board at Emory University and the Add Health study review boards gave ethical approval for this work.

