## Supplemental Table 1 for "Characterization of Trajectories of Physical Activity and Cigarette Smoking from Early Adolescence to Adulthood"

**Supplemental Table 1. Model Selection Criteria for Trajectories of Physical Activity Score and Cigarette Smoking Intensity from Early Adolescence to Adulthood**

| | | | |
|---|---|---|---|
| **Physical Activity** | | | |
| | **AIC** | **BIC** | **Entropy** |
| **1-class model** | -49912.7 | -49833.3 | 1.00 |
| **2-class model** | -49904.7 | -49793.5 | 0.31 |
| **3-class model** | -52101.1 | -51958.2 | 0.60 |
| **4-class model** | -51717.3 | -51542.7 | 0.18 |
| **Optimal Model Class Membership** | | | |
| | **Class 1** | **Class 2** | **Class 3** |
| **N (%)** | 1067 (5) | 14257 (69) | 5410 (26) |
| **Distribution of posterior probability of class membership** | **1st Quartile** | **Mean** | |
| Class 1 | 0.62 | 0.78 | |
| Class 2 | 0.77 | 0.85 | |
| Class 3 | 0.60 | 0.72 | |
| **Log(Past 30-day cigarette smoking intensity)** | | | |
| | **AIC** | **BIC** | **Entropy** |
| **1-class model** | 304046.8 | 304126.1 | 1.00 |
| **2-class model** | 304054.8 | 304165.9 | 0.46 |
| **3-class model** | 284475.4 | 284618.3 | 0.90 |
| **4-class model** | 284483.4 | 284658.1 | 0.59 |
| **3-class Model Class Membership (Optimal Model)** | | | |
| | **Class 1** | **Class 2** | **Class 3** |
| **N (%)** | 14939 (72) | 2357 (11) | 3393 (16) |
| **3-class model probability of class membership** | **1st Quartile** | **Mean** | |
| Class 1 | 0.99 | 0.97 | |
| Class 2 | 0.89 | 0.92 | |
| Class 3 | 0.90 | 0.92 | |
